# Machine Learning-based prediction of Early Neurological Deterioration after Thrombolysis in Acute Ischemic Stroke

**DOI:** 10.1101/2023.02.22.23286330

**Authors:** Yuan Gao, Ce Zong, Hongbing Liu, Ke Zhang, Hongxun Yang, Anran Wang, Yunchao Wang, Yapeng Li, Kai Liu, Yusheng Li, Jing Yang, Bo Song, Yuming Xu

**Affiliations:** Department of Neurology, the First Affiliated Hospital of Zhengzhou University, Zhengzhou, Henan, China

**Keywords:** early neurological deterioration, machine learning, thrombolysis, acute ischemic stroke

## Abstract

**Background:** Early neurological deterioration (END) after thrombolysis in acute ischemic stroke (AIS) cannot be ignored. Our aim is to establish an interpretable machine learning (ML) prediction model for clinical practice.

**Methods:** Patients in this study were enrolled from a prospective, multi-center, web-based registry database. Demographic information, treatment information and laboratory tests were collected. END was defined as an increase of ⩾2 points in total National Institutes of Health Stroke Scale (NIHSS) score within 24 hours after thrombolysis. Eight ML models were trained in the training set (70%) and the tuned models were evaluated in the test set (30%) by calculating the area under the curve (AUC), sensitivity, specificity, accuracy, and F1 scores. Calibration curves were plotted and brier scores were calculated. The SHapley Additive exPlanations (SHAP) analysis and web application were developed for interpretation and practice.

**Results:** A total of 1956 patients were included in the analysis. Of these, 305 patients (15.6%) experienced END. We used logistic regression to identify six important variables: hemoglobin, white blood cell count, the ratio of lymphocytes to monocytes (LMR), thrombin time, onset to treatment time, and prothrombin time. In the test set, the results showed that the Extreme gradient boosting (XGB) model (AUC 0.754, accuracy 0.722, sensitivity 0.723, specificity 0.720, F1 score 0.451) exhibited relatively good performance. Calibration curves showed good agreement between the predicted and true probabilities of the XGB (brier score=0.016) model. We further developed a web application based on it by entering the values of the variables (https://ce-bit123-ml-app1-13tuat.streamlit.app/).

**Conclusions:** Through the identification of critical features and ML algorithms, we developed a web application to help clinicians identify high-risk of END after thrombolysis in AIS patients more quickly, easily and accurately as well as making timely clinical decisions.

## INTRODUCTION

Ischemic stroke is considered to be one of the most lethal and disabling diseases,^1, 2^ and thrombolytic therapy is widely recommended for patients with acute ischemic stroke (AIS).^3^ The use of alteplase is generally acknowledged to be a particularly effective treatment.^4, 5^ However, early neurological deterioration (END) after thrombolysis cannot be ignored. Studies showed that the incidence of END within 24 hours after thrombolysis was 5.8%-20%,^6, 7^ resulting in an adverse prognosis for AIS patients.^8^ Previous studies demonstrated that a number of risk factors such as blood glucose level, National Institutes of Health Stroke Scale (NIHSS) score on admission, blood pressure level and proximal artery occlusion were associated with the occurrence of END after thrombolysis.^7, 9-11^ However, a large-scale and multi-center study to further define the factors associated with the risk of END after thrombolysis was still insufficient, and the disclosure of these risk factors could not be appropriately applied to clinical decision making. In recent years, machine learning (ML) has gradually been widely used in the medical field, and several prognostic and diagnostic models have been developed for ischemic stroke patients.^12, 13^ Optimization of models by fitting machine learning to big data has shown good predictive performance. Such ML based modeling solutions can help clinicians to better individualize their treatments. Based on a prospective, multi-center and large-scale registry study, our aims were to identify risk factors associated with the occurrence of END within 24 hours after thrombolysis in AIS patients as well as establishing an interpretable ML prediction model for clinical practice.

## METHODS

### Data Availability Statement

Data are available upon reasonable request.

### Study Population

Patients in this study were enrolled from a prospective, multi-center, web-based registry study (ChiCTR2100045258) from January 2021 to January 2022.^14^ This registry study enrolled patients with acute ischemic stroke who were admitted to regional stroke centers. Throughout the entire study, information on patients demographics, treatments and prognosis is collected in a standardized manner, and the integrity of the data is monitored monthly by dedicated quality control staff.

In this study, the inclusion criteria were as follows: 1) Diagnosed with acute ischemic stroke by diffusion weighted imaging (DWI); 2) Age⩾18 years; 3) Received thrombolytic therapy; 4) Signed informed consent. Exclusion criteria were as follows: 1) Those who received urokinase thrombolysis; 2) Those with onset to admission time >4.5 hours; 3) Those who received endovascular therapy after thrombolysis; 4) Patients with hematologic disease, severe renal or hepatic disease, cancer, or a history of surgery; 5) Patients with incomplete clinical data. This study was approved by the Ethics Committee of the First Affiliated Hospital of Zhengzhou University.

### Data collection

Demographic information (including age, gender, previous disease history, etc.), treatments information and laboratory tests (including blood count, lipids, liver and kidney function, coagulation function, etc.) were collected. Hypertension was defined as systolic blood pressure (SBP) ⩾ 140 mmHg or diastolic blood pressure (DBP) ⩾ 90 mmHg or taking antihypertensive medication. Hyperglycemia was defined as fasting blood glucose (FBG) >7.0 mmol/L or glycosylated hemoglobin (HbA1c)⩾6.5% or taking glucose-lowering drugs. History of smoking was defined as current smoking or having quited smoking within 6 months. History of drinking was defined as current alcohol use or abstinence within 6 months. Neutrophil to lymphocyte ratio (NLR) was defined as the ratio of neutrophils to lymphocytes. Lymphocyte to monocyte ratio (LMR) was defined as the ratio of lymphocytes to monocytes. Patients enrolled in each stroke center were assessed of the NIHSS score by 2 designated neurologists respectively during consecutive seven days after admission. These neurologists had uniform specialty training, which were blinded to the study. In our study, END was defined as an increase of ⩾ 2 points in total NIHSS score within 24 hours after thrombolysis.^7, 15^

### Data processing and feature selection

We finally selected 40 variables referring to previous literature and clinical practice, which included 8 demographic characteristics, 7 previous disease history, 4 treatments and 21 laboratory indicators. These variables were also common and readily available in clinical practice. We performed univariate analysis for these variables and multivariate logistic regression (LR) analysis for those variables with p<0.05. Correlation analysis was conducted to determine the magnitude of correlation between variables. The final variables for inclusion in the ML model were determined based on the multivariate LR results.

### Model construction and evaluation

In this study, we used eight ML models: LR, Support Vector Machine(SVM), Decision tree(DT),Random Forest (RF), Gradient Boosting Machine (GBM), Extreme gradient boosting (XGB), Light Gradient Boosting Machine (LightGBM) and Categorical Boosting (CatBoost). The dataset was randomly divided into two sets: the training set (70%) for model training and hyperparameter optimization, and the test set (30%) for validating the predictive power of the models. In the training set, we used grid search to perform hyperparameter optimization to determine the best model hyperparameters, and then applied the tuned model to the test set to evaluate the predictive value of the models by calculating the area under the curve (AUC), sensitivity, specificity, accuracy, and F1 scores. Calibration curves were plotted and brier scores were calculated to determine the consistency between the predicted and true values of the models. Ultimately, we identified the best performing model for the subsequent analysis.

### Model explanation and application

To address the “black boxes” issues of ML models that were difficult to explain the results directly, we interpreted the model by means of the SHapley Additive exPlanations (SHAP) analysis and visualized the SHAP values to determine the ranking of feature importance in the optimal model and the impact on the outcome events. In addition, we developed a web application based on the optimal model to aid clinical practice.

### Statistical analysis

All data were analyzed and visualized using R version 4.1.2 and python 3. Missing values >20% and outliers were excluded, and missing values were filled using the mean for continuous variables and the plural for categorical variables. In univariate analysis, continuous variables were presented using mean ± standard deviation or Student’s t-test or Mann-Whitney U test. Categorical variables were presented using proportions by chi-square test. p<0.05 was considered significant. The data were randomly divided into training and test sets in a ratio of 7:3. When training the model, all data were standardized. We optimized the hyperparameters using a 10-fold cross-validation in the training set by GridSearch CV and calculated the evaluation metrics (including the AUC, sensitivity, specificity, accuracy, F1 scores and brier scores) of the tuned models in the validation set. When evaluating the model performance, we used AUC and F1 scores to assess model distinction and brier scores to assess model calibration. After selecting the optimal model, the model was interpreted by SHAP value analysis and a web application was developed with the streamlit platform to guide clinical decision making.

## RESULTS

### Baseline

A total of 2,353 ischemic stroke patients were received thrombolytic therapy from January 2021 to January 2022, of which, 122 were thrombolized with urokinase, 44 had onset to admission time exceeding 4.5 hours, 20 underwent procedures such as mechanical thrombectomy, 102 suffered from hematological system or severe renal and hepatic diseases, etc. and 109 had incomplete data. Finally, a total of 1956 patients were included in the final analysis. Of these, 305 patients (15.6%) experienced END. the mean age was 66.3 ± 11.4 years old, and 755 patients (38.6%) were female. Additional clinical information was detailed in **Table 1.**

**Table 1.**
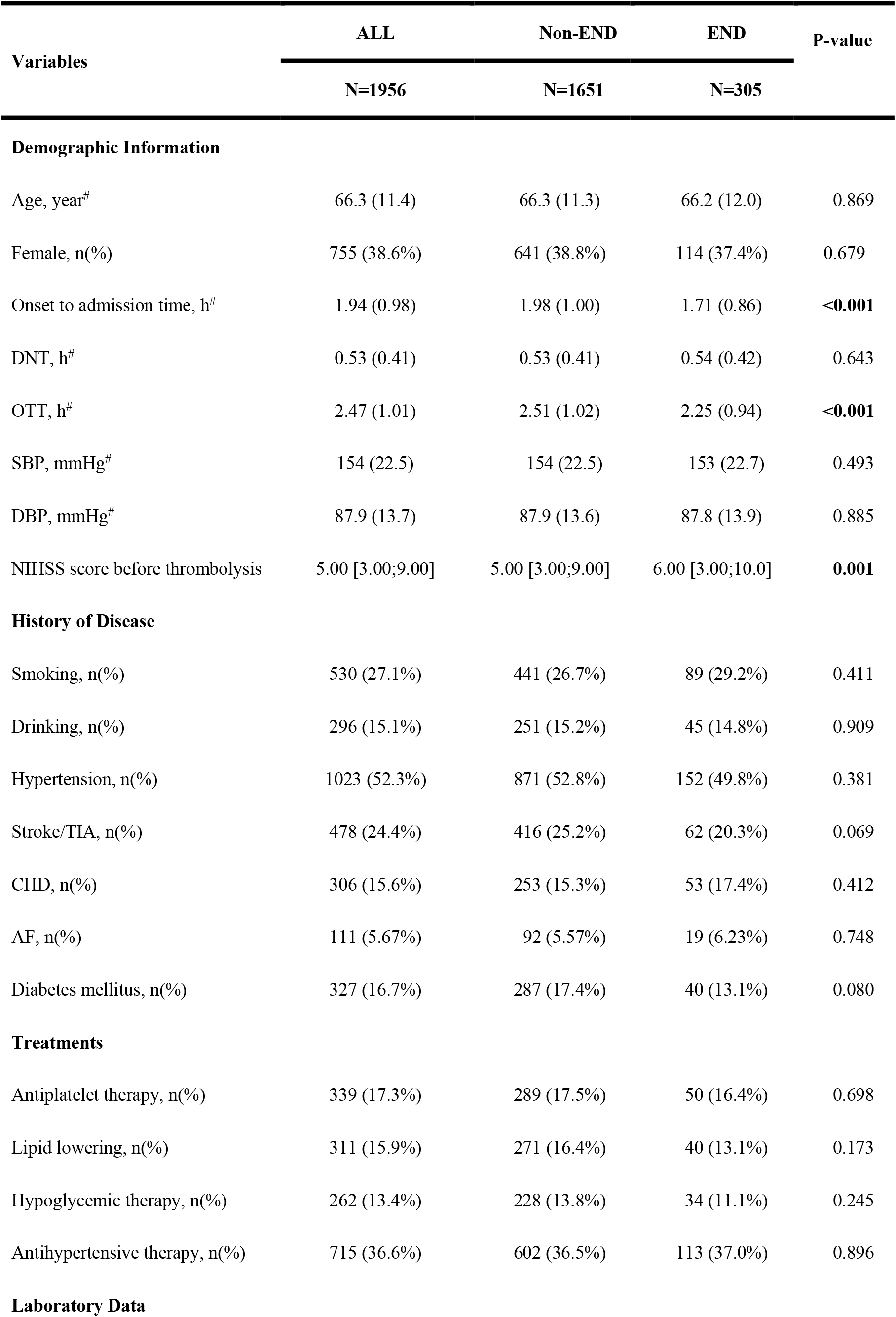
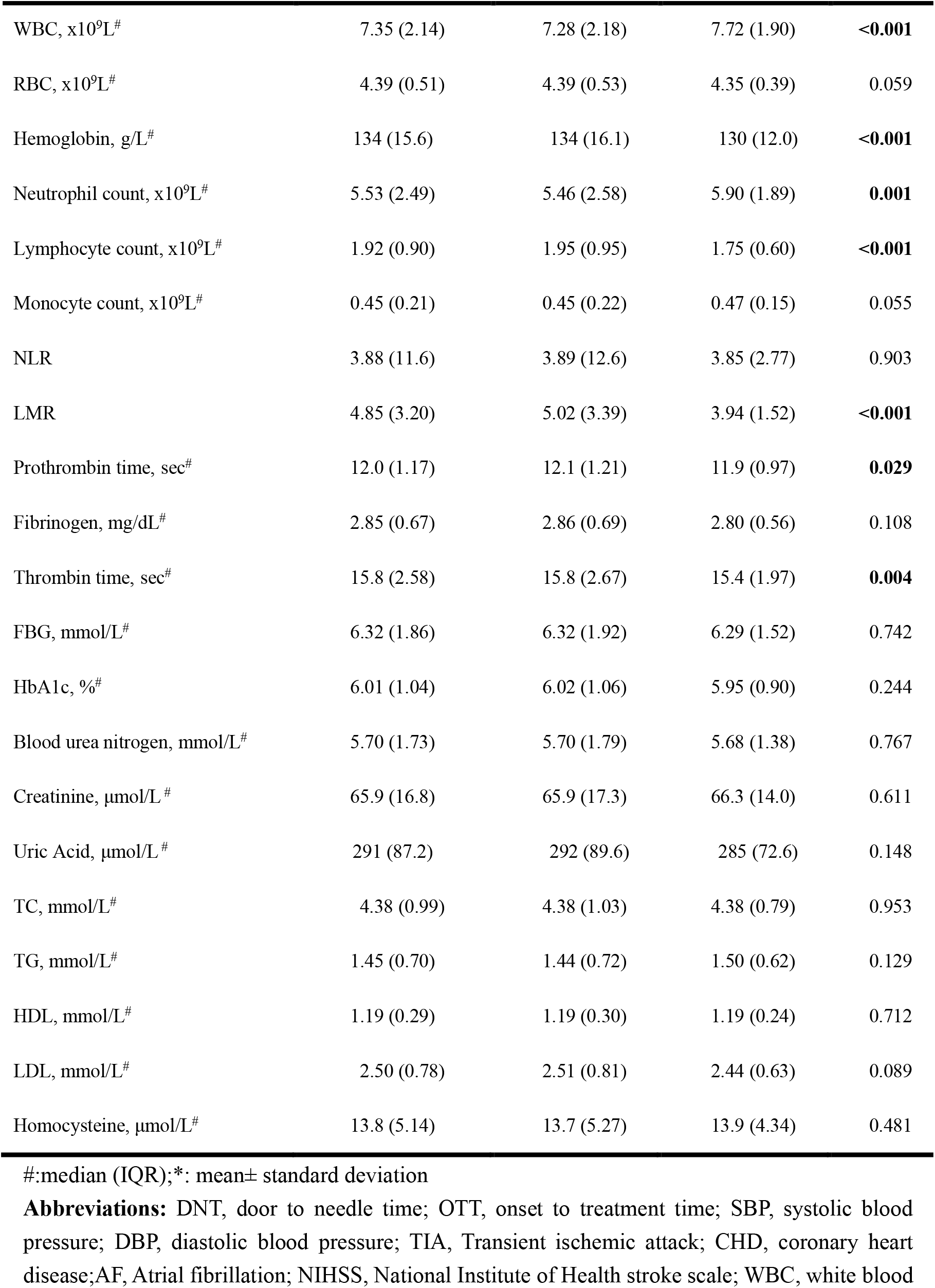
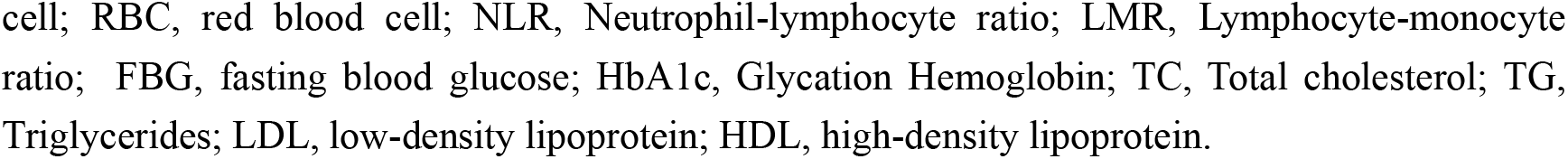
Comparison of clinical characteristics between ischemic stroke patients with END and Non-END occurring within 24 hours after thrombolysis.

| Variables | ALL<br>N=1956 | Non-END<br>N=1651 | END<br>N=305 | P-value |
| --- | --- | --- | --- | --- |
| **Demographic Information** |  |  |  |  |
| Age, year<sup>#</sup> | 66.3 (11.4) | 66.3 (11.3) | 66.2 (12.0) | 0.869 |
| Female, n(%) | 755 (38.6%) | 641 (38.8%) | 114 (37.4%) | 0.679 |
| Onset to admission time, h<sup>#</sup> | 1.94 (0.98) | 1.98 (1.00) | 1.71 (0.86) | **<0.001** |
| DNT, h<sup>#</sup> | 0.53 (0.41) | 0.53 (0.41) | 0.54 (0.42) | 0.643 |
| OTT, h<sup>#</sup> | 2.47 (1.01) | 2.51 (1.02) | 2.25 (0.94) | **<0.001** |
| SBP, mmHg<sup>#</sup> | 154 (22.5) | 154 (22.5) | 153 (22.7) | 0.493 |
| DBP, mmHg<sup>#</sup> | 87.9 (13.7) | 87.9 (13.6) | 87.8 (13.9) | 0.885 |
| NIHSS score before thrombolysis | 5.00 [3.00;9.00] | 5.00 [3.00;9.00] | 6.00 [3.00;10.0] | **0.001** |
| **History of Disease** |  |  |  |  |
| Smoking, n(%) | 530 (27.1%) | 441 (26.7%) | 89 (29.2%) | 0.411 |
| Drinking, n(%) | 296 (15.1%) | 251 (15.2%) | 45 (14.8%) | 0.909 |
| Hypertension, n(%) | 1023 (52.3%) | 871 (52.8%) | 152 (49.8%) | 0.381 |
| Stroke/TIA, n(%) | 478 (24.4%) | 416 (25.2%) | 62 (20.3%) | 0.069 |
| CHD, n(%) | 306 (15.6%) | 253 (15.3%) | 53 (17.4%) | 0.412 |
| AF, n(%) | 111 (5.67%) | 92 (5.57%) | 19 (6.23%) | 0.748 |
| Diabetes mellitus, n(%) | 327 (16.7%) | 287 (17.4%) | 40 (13.1%) | 0.080 |
| **Treatments** |  |  |  |  |
| Antiplatelet therapy, n(%) | 339 (17.3%) | 289 (17.5%) | 50 (16.4%) | 0.698 |
| Lipid lowering, n(%) | 311 (15.9%) | 271 (16.4%) | 40 (13.1%) | 0.173 |
| Hypoglycemic therapy, n(%) | 262 (13.4%) | 228 (13.8%) | 34 (11.1%) | 0.245 |
| Antihypertensive therapy, n(%) | 715 (36.6%) | 602 (36.5%) | 113 (37.0%) | 0.896 |
| **Laboratory Data** |  |  |  |  |

| Variables | ALL | Non-END | END | P-value |
| --- | --- | --- | --- | --- |
|  | N=1956 | N=1651 | N=305 |  |
| WBC, x10<sup>9</sup>L<sup>#</sup> | 7.35 (2.14) | 7.28 (2.18) | 7.72 (1.90) | <0.001 |
| RBC, x10<sup>9</sup>L<sup>#</sup> | 4.39 (0.51) | 4.39 (0.53) | 4.35 (0.39) | 0.059 |
| Hemoglobin, g/L<sup>#</sup> | 134 (15.6) | 134 (16.1) | 130 (12.0) | <0.001 |
| Neutrophil count, x10<sup>9</sup>L<sup>#</sup> | 5.53 (2.49) | 5.46 (2.58) | 5.90 (1.89) | 0.001 |
| Lymphocyte count, x10<sup>9</sup>L<sup>#</sup> | 1.92 (0.90) | 1.95 (0.95) | 1.75 (0.60) | <0.001 |
| Monocyte count, x10<sup>9</sup>L<sup>#</sup> | 0.45 (0.21) | 0.45 (0.22) | 0.47 (0.15) | 0.055 |
| NLR | 3.88 (11.6) | 3.89 (12.6) | 3.85 (2.77) | 0.903 |
| LMR | 4.85 (3.20) | 5.02 (3.39) | 3.94 (1.52) | <0.001 |
| Prothrombin time, sec<sup>#</sup> | 12.0 (1.17) | 12.1 (1.21) | 11.9 (0.97) | 0.029 |
| Fibrinogen, mg/dL<sup>#</sup> | 2.85 (0.67) | 2.86 (0.69) | 2.80 (0.56) | 0.108 |
| Thrombin time, sec<sup>#</sup> | 15.8 (2.58) | 15.8 (2.67) | 15.4 (1.97) | 0.004 |
| FBG, mmol/L<sup>#</sup> | 6.32 (1.86) | 6.32 (1.92) | 6.29 (1.52) | 0.742 |
| HbA1c, %<sup>#</sup> | 6.01 (1.04) | 6.02 (1.06) | 5.95 (0.90) | 0.244 |
| Blood urea nitrogen, mmol/L<sup>#</sup> | 5.70 (1.73) | 5.70 (1.79) | 5.68 (1.38) | 0.767 |
| Creatinine, μmol/L<sup>#</sup> | 65.9 (16.8) | 65.9 (17.3) | 66.3 (14.0) | 0.611 |
| Uric Acid, μmol/L<sup>#</sup> | 291 (87.2) | 292 (89.6) | 285 (72.6) | 0.148 |
| TC, mmol/L<sup>#</sup> | 4.38 (0.99) | 4.38 (1.03) | 4.38 (0.79) | 0.953 |
| TG, mmol/L<sup>#</sup> | 1.45 (0.70) | 1.44 (0.72) | 1.50 (0.62) | 0.129 |
| HDL, mmol/L<sup>#</sup> | 1.19 (0.29) | 1.19 (0.30) | 1.19 (0.24) | 0.712 |
| LDL, mmol/L<sup>#</sup> | 2.50 (0.78) | 2.51 (0.81) | 2.44 (0.63) | 0.089 |
| Homocysteine, μmol/L<sup>#</sup> | 13.8 (5.14) | 13.7 (5.27) | 13.9 (4.34) | 0.481 |

<sup>#</sup>:median (IQR);\*: mean± standard deviation
**Abbreviations:** DNT, door to needle time; OTT, onset to treatment time; SBP, systolic blood pressure; DBP, diastolic blood pressure; TIA, Transient ischemic attack; CHD, coronary heart disease; AF, Atrial fibrillation; NIHSS, National Institute of Health stroke scale; WBC, white blood
cell; RBC, red blood cell; NLR, Neutrophil-lymphocyte ratio; LMR, Lymphocyte-monocyte ratio; FBG, fasting blood glucose; HbA1c, Glycation Hemoglobin; TC, Total cholesterol; TG, Triglycerides; LDL, low-density lipoprotein; HDL, high-density lipoprotein.

### Feature selection and model development

We compared the clinical characteristics of patients who developed END versus Non-END during 24 hours after thrombolysis. The results showed shorter onset to admission time (P <0.001), shorter onset to treatment time (OTT) (P <0.001), higher NIHSS score before thrombolysis (P =0.001), higher white blood cell (WBC) counts (P <0.001), hemoglobin (P <0.001) and neutrophil count (P =0.001), lower lymphocyte counts (P<0.001) and LMR (P <0.001), and shorter prothrombin time (P = 0.029) and thrombin time (P = 0.004) in patients in the END group compared with the Non-END group **(Table 1).**

The correlation coefficients between these variables with P<0.05 in the univariate analysis were then calculated and a heat map of the correlation coefficients was performed **(Figure S1).** Among them, onset to admission time and OTT, WBC and neutrophil count, lymphocyte count and LMR were highly correlated. Based on previous literature and the practicality of clinical work, we finally selected OTT, WBC and LMR and performed multivariate LR analysis together with other variables. The final results showed that OTT (adjusted OR 0.74, 95%CI 0.65-0.84, P<0.001), hemoglobin (adjusted OR 0.98, 95%CI 0.97-0.99, P<0.001), thrombin time (adjusted OR 0.95, 95%CI 0.90-1, P=0.041), prothrombin time (adjusted OR 0.86, 95%CI 0.76-0.96, P=0.007) and LMR (adjusted OR 0.80, 95%CI 0.74-0.86, P<0.001) were negatively associated with the occurrence of END, while WBC (adjusted OR 1.08, 95%CI 1.02-1.14, P=0.013) was positively correlated independently **(Table 2).** We finally selected the above six variables for the construction of the ML model.

**Table 2.** Multivariate analysis of risk factors associated with the occurrence of END within 72 hours after thrombolysis.

| Variables | Adjusted OR* | 95%CI | P-value |
| --- | --- | --- | --- |
| OTT | 0.74 | 0.65-0.84 | <0.001 |
| WBC | 1.08 | 1.02-1.14 | 0.013 |
| Hemoglobin | 0.98 | 0.97-0.99 | <0.001 |
| Thrombin time | 0.95 | 0.90-1.00 | 0.041 |
| Prothrombin time | 0.86 | 0.76-0.96 | 0.007 |
| LMR | 0.80 | 0.74-0.86 | <0.001 |

**Abbreviations:** END, early neurological deterioration; WBC, White blood cell; OTT, onset to treatment time; LMR, Lymphocyte-monocyte ratio; OR, odds ratio; CI: confidence interval; P-value <0.05 was considered meaningful.
\*Adjusted for NIHSS score before thrombolysis.

### Model performance and evaluation

Of all 1956 patients, 1369 patients (70%) were assigned to the training set while 587 patients (30%) were in the test set. We performed hyperparameter optimization of the models in the training set and used the best trained models in the test set, The results showed that the XGB model (AUC 0.754, accuracy 0.722, sensitivity 0.723, specificity 0.720, F1 score 0.451) and the CatBoost model (AUC 0.752, accuracy 0.739, sensitivity 0.753, specificity 0.667, F1 score 0.451) exhibited relatively good performance **(Table 3, Figure 1A).**

**Figure 1.**
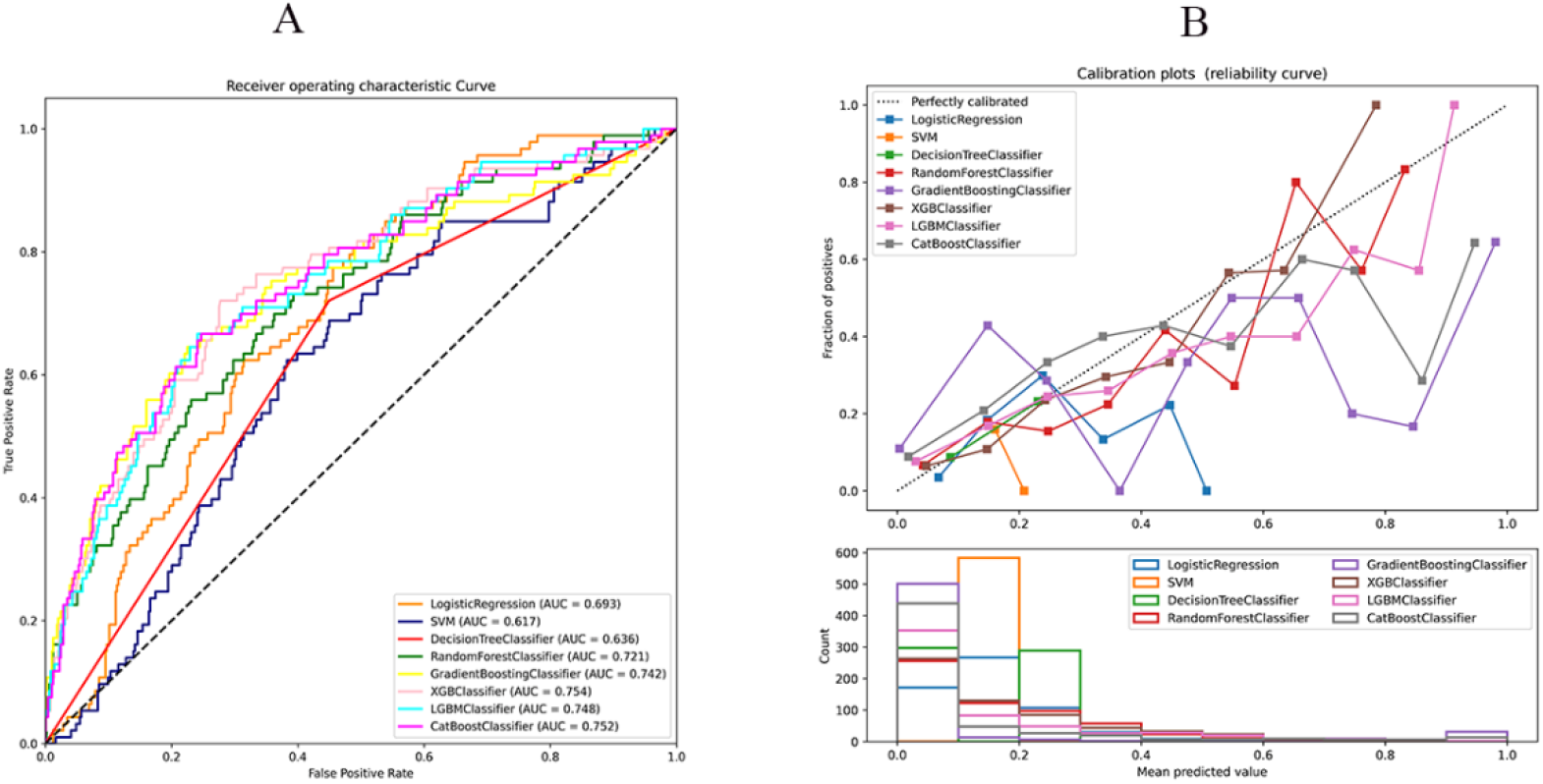
(A)The area under the curves (ROCs) in the test set between different models. (B)Calibration curves in the test set between different models.

**Table 3.** Comparison results of evaluation indexes between different models.

| Model name | Train_AUC | Brier score | Test_AUC | Accuracy | Specificity | Sensitivity | F1 |
| --- | --- | --- | --- | --- | --- | --- | --- |
| LR | 0.709 | 0.129 | 0.693 | 0.581 | 0.545 | 0.774 | 0.369 |
| SVM | 0.614 | 0.132 | 0.617 | 0.618 | 0.617 | 0.624 | 0.341 |
| DT | 0.637 | 0.128 | 0.636 | 0.578 | 0.551 | 0.720 | 0.351 |
| RF | 0.997 | 0.119 | 0.721 | 0.627 | 0.607 | 0.731 | 0.383 |
| GBM | 1 | 0.139 | 0.742 | 0.755 | 0.775 | 0.645 | 0.455 |
| XGB | 0.894 | 0.116 | 0.754 | 0.722 | 0.723 | 0.720 | 0.451 |
| LGBM | 0.998 | 0.120 | 0.748 | 0.744 | 0.759 | 0.667 | 0.453 |
| CatBoost | 1 | 0.125 | 0.752 | 0.739 | 0.753 | 0.667 | 0.448 |

**Abbreviations:** LR, Logistic Regression; SVM, Support Vector Machine; DT, Decision tree; RF, Random Forest; GBM, Gradient Boosting Machine; XGB, Extreme gradient boosting; LightGBM, Light Gradient Boosting Machine; CatBoost, Categorical Boosting.

In addition, the calibration curves showed good agreement between the predicted and true probabilities of the XGB (brier score=0.016) and CatBoost (brier score=0.125) models **(Table 3, Figure 1B).** We finally selected the XGB model as the final explanatory model and established the web application based on it in terms of the AUC and F1 score of the models in the test set.

### Model explanation and application

We investigated the order of importance of the variables to the XGB model by calculating the mean absolute SHAP values, and the results showed that the six were in descending order of importance to the model: LMR, hemoglobin, WBC, thrombin time, OTT, and prothrombin time **(Figure 2A).** Impacts of the variables on the occurrence of END within 24 hours after thrombolysis were showed, and the results revealed that LMR, hemoglobin, thrombin time, OTT, and prothrombin time had a negative effect on END, whereas WBC counts had a positive effect on END (**Figure 2B**).

**Figure 2.**
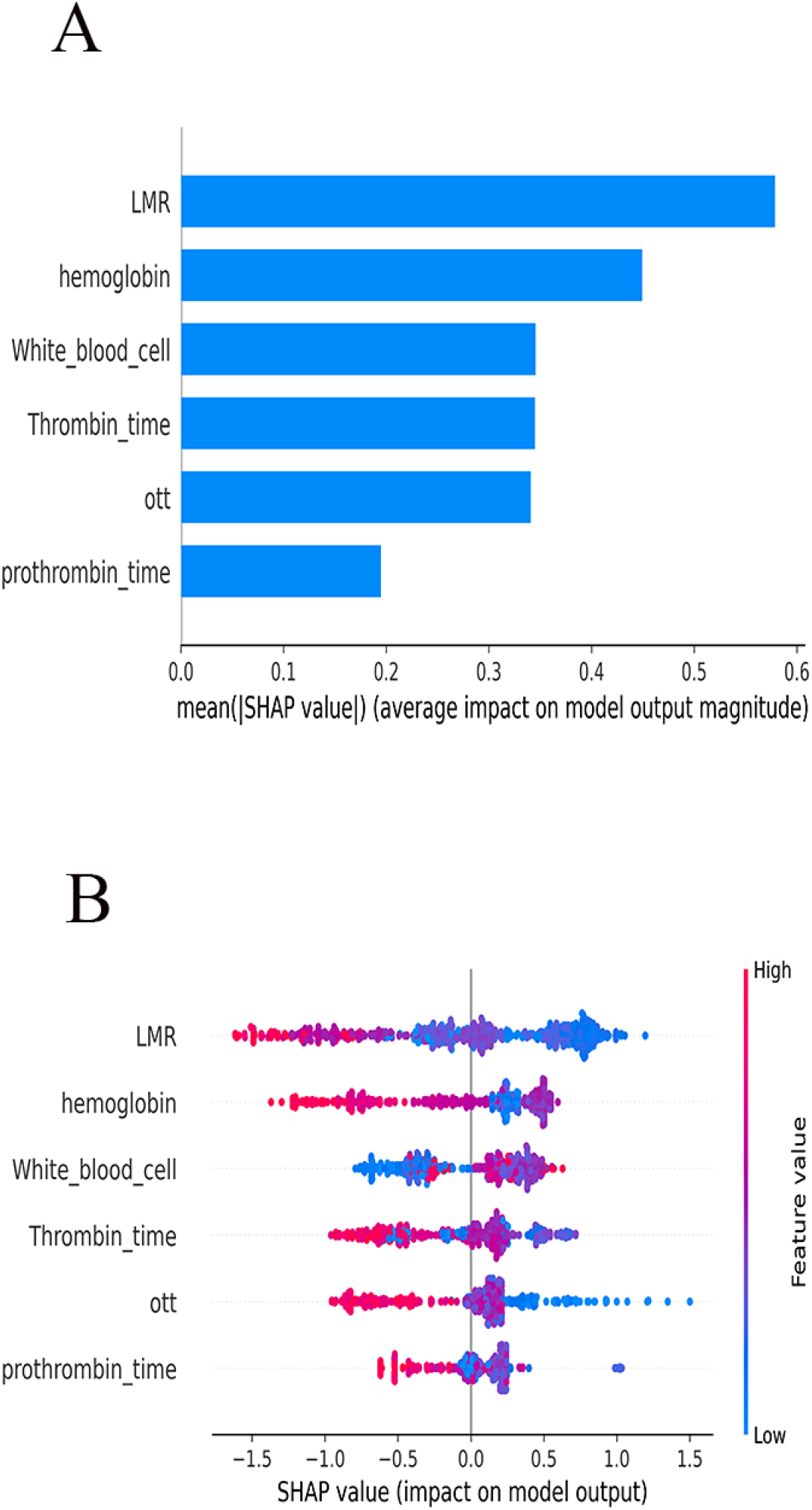
SHAP summary plot of the importance of variables based on XGB model for predicting END (A, B).

We further developed a web application based on the XGB model to predict the risk of END within 24 hours of thrombolysis in individual ischemic stroke patients by entering the values of the variables **(Figure 3)**. (https://ce-bit123-ml-app1-13tuat.streamlit.app/)

**Figure 3.**
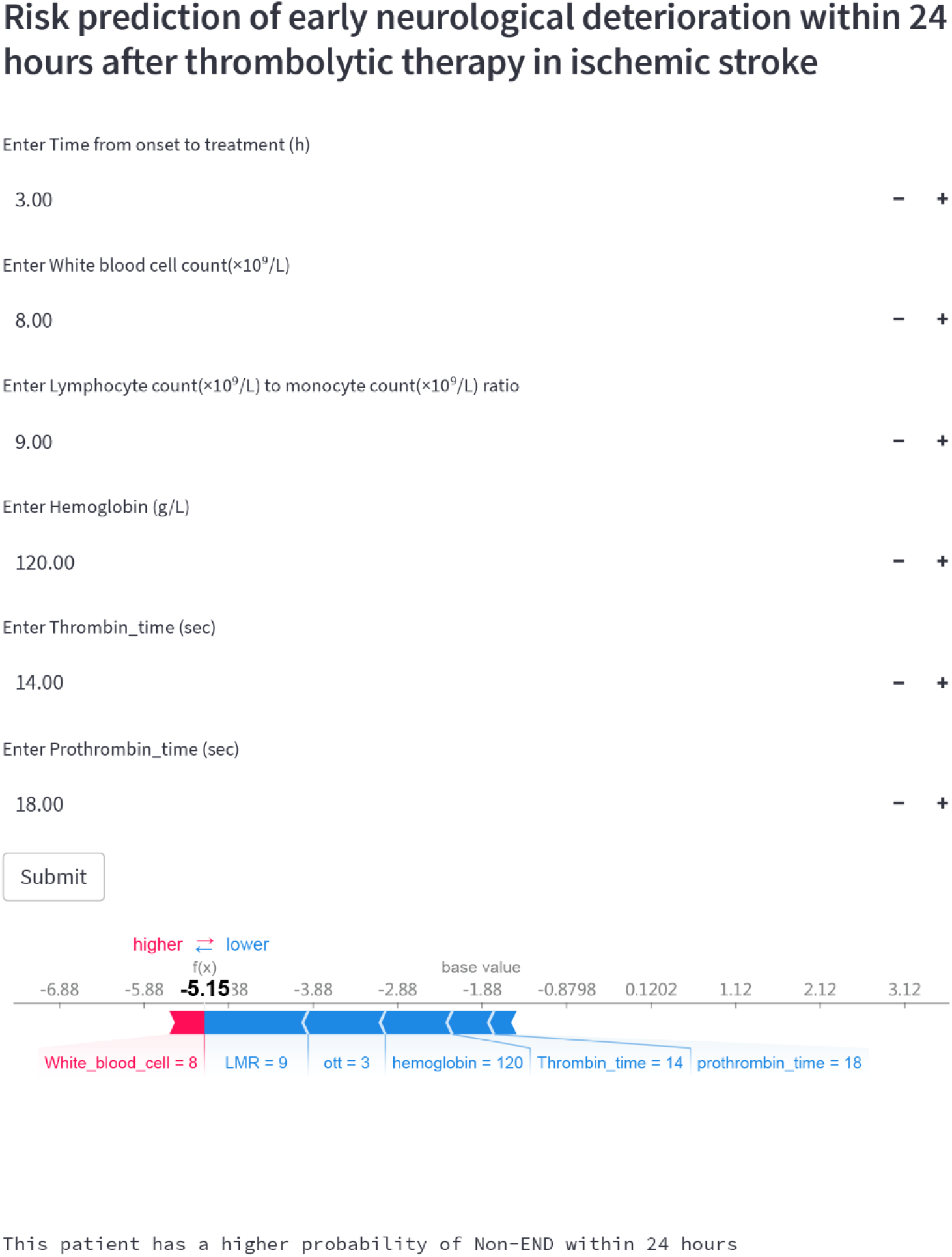
Web app based on XGB model for predicting the risk of END in ischemic stroke patients within 72 hours after thrombolysis. (https://ce-bit123-ml-app1-13tuat.streamlit.app/)

## DISCUSSION

In this study, we performed feature selection by LR, and finally picked these six variables: LMR, hemoglobin, WBC, thrombin time, OTT, and prothrombin time. We applied eight machine learning algorithms to construct and validate a predictive model for the risk of END within 24 hours after thrombolysis for AIS patients in a large-scale and multi-center study in domestic population, and discovered that several machine models (RF, GBM, XGB, LGBM and CatBoost) showed higher predictive power than the conventional logistic regression model, in which the XGB model performed the best. Further, we developed a web application based on the XGB model using the streamlit platform architecture to enhance the clinical utility by facilitating risk prediction for clinicians.

A growing number of studies have demonstrated the benefit of alteplase thrombolysis in patients with AIS within the time window.^5, 16^ However, it does not mean that thrombolysis is absolutely safe, and an inescapable complication concerns the occurrence of END, which may be caused by intracranial hemorrhage and malignant edema, etc.^16^ Therefore, studies on the risk prediction of END after thrombolysis are particularly valuable. Previously, Chung CC et al developed an artificial neural network-based model to help predict the major neurological improvements within 24 hours and 3-month prognosis after thrombolysis in 196 patients with AIS treated with intravenous thrombolysis in a Taiwan hospital.^17^ Seners P et al constructed a risk score to predict presumed ischemic-derived END in a multi-center and retrospective cohort study including 729 consecutive patients with mild stroke (NIHSS score ≤5) and large vessel occlusion (LVO).^18^ However, there were problems with poor model selection, limited application and small samples in these researches, while our study was conducted in a large-scale, multi-center and domestic population, using more advanced ML algorithms to construct models and SHAP value analysis as well as a web application to make the findings more applicable. Meanwhile, our study provided new insights into the etiological mechanisms of END.

In our interpretable model, in addition to identifying significant indicators, we also clarified the impact of these factors on END and the magnitude of feature importance. These findings would assist us in gaining a deeper understanding of the mechanisms underlying the occurrence of END. Our results suggested that inflammatory markers (LMR and WBC) probably played an influential role in the occurrence of END. In fact, previous studies showed that lower LMR was associated with poor clinical prognosis at 3 months after thrombolysis in patients with AIS.^19^ Mao X et al discovered that LMR was an independent predictor of progressive infarction in patients with AIS (especially the large atherosclerotic type) by retrospectively analyzing 477 AIS patients within 48 hours of onset.^20^ While WBC counts were proved to be related to ischemic stroke and cardiovascular disease.^21^ Furlan JC et al found that higher WBC on admission was an independently predictive factor for stroke severity on admission, greater disability at discharge as well as 30-day mortality in a study of 8829 AIS patients from the Canadian Stroke Network registry.^22^ Immune inflammatory responses are involved in the development of ischemic stroke,^23, 24^ in which lymphocytes are neuroprotective while monocytes can disrupt the blood-brain barrier by secreting pro-inflammatory factors, further aggravating brain tissue damage.^19, 23^ WBC counts can adhere to the endothelium and damage it through toxic oxidants and protein hydrolases.^25^ Elevated WBC counts also increase pro-inflammatory factors that participate in the clotting process and consequently predispose to thrombus formation.^26^ In addition, leukocyte aggregation and activation are amplified by the action of thrombolytic drugs, causing persistent tissue ischemia.^25, 27^ Therefore, both reduced LMR and elevated WBC counts may indicate a larger immune inflammatory response during ischemic stroke, and reperfusion after thrombolysis may further lead to increased tissue damage in the inflammatory response and greater susceptibility to END. Attention to changes in inflammatory markers in the early stages of stroke may be extremely important for the prevention of END after thrombolysis.

In our study, we found that lower hemoglobin was also a critical factor associated with the occurrence of END. Previous studies showed that lower hemoglobin during hospitalization was linked to larger and faster growing infarct size at baseline, even when hemoglobin was not reduced to the defined level of anemia.^28^ Reduced hemoglobin represents a decrease in oxygen-carrying capacity of the blood, which can affect the self-regulation of brain tissue resulting in infarct enlargement and the development of END.^29, 30^ Meanwhile, our results suggested that OTT was also significant. In the univariate analysis, DNT was not significantly different between the END and Non-END groups (p=0.643), while the time from onset to admission was shorter in the END group. Considering that the included patients had a lower admission NIHSS score (5.00 [3.00-9.00]), these patients were probably in the early stages of onset and had more potential for further exacerbation, who still should be brought to our enough attention.

Meanwhile, it remained worth noting the effect of coagulation (thrombin time and prothrombin time) on END. Studies showed that the use of alteplase could improve the coagulation function of ischemic stroke patients, and activate fibrinolytic enzymes to destroy thrombus as well as reducing thrombus formation to achieve the effect of thrombolysis.^31^ Reduced thrombin time and prothrombin time implies an increase in clotting activity resulting in greater susceptibility to new thrombus formation. In this case, it could cause thrombus expansion or a new embolic event.^32^ Alternatively, it may cause inadequate thrombus lysis and consequently hemodynamic disturbances that could lead to END.^33^ Therefore, we probably also need to pay attention to the changes in coagulation parameters before and after thrombolysis in patients in order to be alerted to the occurrence of END earlier.

The final part of our study was to develop a web application using the streamlit platform to obtain a prediction of the risk of END occurring within 24 hours of thrombolysis in patients by entering the six variable values. The application would assist clinicians in the risk stratification of patients undergoing thrombolysis, thus allowing earlier identification of high-risk groups and prompt management.

There are some limitations in our study: Firstly, our research was conducted in a Chinese population exclusively, and further research is required to determine whether it is suitable for other population. Secondly, there is also a lack of uniformity in terms of END after thrombolysis, and the difference in the definitions of END will lead to the variation of analysis results. Finally, although we have included 40 variables with reference to previous literature and clinical practice, it remains the case that potentially meaningful variables have not been included in the analysis, which may introduce some information omission.

## CONCLUSION

In this study, we used LR to identify six important variables (including LMR, hemoglobin, WBC, thrombin time, OTT, and prothrombin time) and applied eight ML algorithms to construct a predictive model for the risk of END occurring within 24 hours after thrombolysis in AIS patients. Of all models, the XGB model performed best based on model discrimination and calibration. We performed SHAP analysis based on the XGB model and developed a web application to help clinicians identify high-risk patients more quickly, easily and accurately as well as making timely clinical decisions.

## Data Availability

Data are available upon reasonable request.

## Non-standard Abbreviations and Acronyms

AIS: acute ischemic stroke
END: early neurological deterioration
NIHSS: National Institutes of Health Stroke Scale
ML: machine learning
LMR: lymphocyte to monocyte ratio
SHAP: SHapley Additive exPlanations
OTT: onset to treatment time
DWI: diffusion weighted imaging
AUC: area under the curve
ROC: receiver operating characteristic

## Acknowledgments

We thank all participants. Yuan Gao, Ce Zong, Bo Song, and Yuming Xu conceived and designed the research. Ce Zong, Ke Zhang, Hongxun Yang, Hongbing Liu,Yunchao Wang acquired the data. Ce Zong analyzed and interpreted the data. Yuan Gao, Ce Zong drafted the manuscript. Yapeng Li, Kai Liu, Yusheng Li and Jing Yang made critical revisions to the manuscript. All authors approved the final manuscript.

## REFERENCE

1. SK F. Ischemic Stroke. The American journal of medicine 2021;134:1457–1464.

2. G Y, Y W, Y Z, et al. Rapid health transition in China, 1990-2010: findings from the Global Burden of Disease Study 2010. Lancet (London, England) 2013;381:1987–2015.

3. WJ P, AA R, T A, et al. 2018 Guidelines for the Early Management of Patients With Acute Ischemic Stroke: A Guideline for Healthcare Professionals From the American Heart Association/American Stroke Association. Stroke 2018;49:e46–e110.

4. AL G, AD B, O A. Alteplase and Adjuvant Therapies for Acute Ischemic Stroke. Seminars in neurology 2021;41:16–27.

5. G T, C I, D C. Intravenous thrombolysis for acute ischemic stroke. Diagnostic and interventional imaging 2014;95:1129–1133.

6. Simonsen CZ, Schmitz ML, Madsen MH, et al. Early neurological deterioration after thrombolysis: Clinical and imaging predictors. Int J Stroke 2016;11:776–782.

7. Nair SB, Somarajan D, Pillai RK, Balachandran K, Sathian S. Predictors of Early Neurological Deterioration Following Intravenous Thrombolysis: Difference between Risk Factors for Ischemic and Hemorrhagic Worsening. Ann Indian Acad Neurol 2022;25:627–633.

8. F C, A W, Y J, et al. Early neurological deterioration in acute ischemic stroke patients after intravenous thrombolysis with alteplase predicts poor 3-month functional prognosis - data from the Thrombolysis Implementation and Monitor of Acute Ischemic Stroke in China (TIMS-China). BMC neurology 2022;22:212.

9. L W, Q C, T H, et al. Impact of Stress Hyperglycemia on Early Neurological Deterioration in Acute Ischemic Stroke Patients Treated With Intravenous Thrombolysis. Frontiers in neurology 2022;13:870872.

10. M M, M N, Y O, et al. Early neurological deterioration within 24 hours after intravenous rt-PA therapy for stroke patients: the Stroke Acute Management with Urgent Risk Factor Assessment and Improvement rt-PA Registry. Cerebrovascular diseases (Basel, Switzerland) 2012;34:140–146.

11. YL L, HP Y, DH Q, et al. Multiple hypointense vessels on susceptibility-weighted imaging predict early neurological deterioration in acute ischaemic stroke patients with severe intracranial large artery stenosis or occlusion receiving intravenous thrombolysis. Stroke and vascular neurology 2020;5:361–367.

12. MS J, O J, D K, et al. Interpretable Machine Learning Modeling for Ischemic Stroke Outcome Prediction. Frontiers in neurology 2022;13:884693.

13. WX Y, FF W, YY P, JQ X, MH L, CG Y. Comparison of ischemic stroke diagnosis models based on machine learning. Frontiers in neurology 2022;13:1014346.

14. Liu H, Liu K, Zhang K, et al. Early neurological deterioration in patients with acute ischemic stroke: a prospective multicenter cohort study. Ther Adv Neurol Disord 2023;16:17562864221147743.

15. Zhou Y, Zhong W, Wang A, et al. Hypoperfusion in lenticulostriate arteries territory related to unexplained early neurological deterioration after intravenous thrombolysis. Int J Stroke 2019;14:306–309.

16. P S, G T, C O, JC B. Incidence, causes and predictors of neurological deterioration occurring within 24 h following acute ischaemic stroke: a systematic review with pathophysiological implications. Journal of neurology, neurosurgery, and psychiatry 2015;86:87–94.

17. CC C, CT H, YH H, et al. Predicting major neurologic improvement and long-term outcome after thrombolysis using artificial neural networks. Journal of the neurological sciences 2020;410:116667.

18. P S, W BH, B L, et al. Prediction of Early Neurological Deterioration in Individuals With Minor Stroke and Large Vessel Occlusion Intended for Intravenous Thrombolysis Alone. JAMA neurology 2021;78:321–328.

19. H R, L H, H L, L W, X L, Y G. Decreased Lymphocyte-to-Monocyte Ratio Predicts Poor Prognosis of Acute Ischemic Stroke Treated with Thrombolysis. Medical science monitor : international medical journal of experimental and clinical research 2017;23:5826–5833.

20. X M, Q Y, Y L, et al. Lymphocyte-to-Monocyte Ratio Is Independently Associated with Progressive Infarction in Patients with Acute Ischemic Stroke. BioMed research international 2022;2022:2290524.

21. CD L, AR F, FJ N, LE C, E S, DA W. White blood cell count and incidence of coronary heart disease and ischemic stroke and mortality from cardiovascular disease in African-American and White men and women: atherosclerosis risk in communities study. American journal of epidemiology 2001;154:758–764.

22. JC F, MD V, J F, FL S. White blood cell count is an independent predictor of outcomes after acute ischaemic stroke. European journal of neurology 2014;21:215–222.

23. R M, C A, O T, et al. Stroke and the immune system: from pathophysiology to new therapeutic strategies. The Lancet Neurology 2011;10:471–480.

24. T L, A L. Immunity in Stroke: The Next Frontier. Thrombosis and haemostasis 2022;122:1454–1460.

25. E E, DE H, U B, A M, JA D. Leukocytes and the risk of ischemic diseases. JAMA 1987;257:2318–2324.

26. Cermak J, Key NS, Bach RR, Balla J, Jacob HS, Vercellotti GM. C-reactive protein induces human peripheral blood monocytes to synthesize tissue factor. Blood 1993;82:513–520.

27. Villemure C, Bushnell MC. Mood influences supraspinal pain processing separately from attention. J Neurosci 2009;29:705–715.

28. S B, R B, M N, et al. Association of anemia and hemoglobin decrease during acute stroke treatment with infarct growth and clinical outcome. PloS one 2018;13:e0203535.

29. WT K, O W, EM A, et al. Lower hemoglobin correlates with larger stroke volumes in acute ischemic stroke. Cerebrovascular diseases extra 2011;1:44–53.

30. L K, C H, M S, et al. Loss of Penumbra by Impaired Oxygen Supply? Decreasing Hemoglobin Levels Predict Infarct Growth after Acute Ischemic Stroke: Stroke: Relevant Impact of Hemoglobin, Hematocrit and Transfusion (STRAIGHT) - An Observational Study. Cerebrovascular diseases extra 2012;2:99–107.

31. HY Z, GT Y, HF Z, WH W. Effect of Alteplase Thrombolysis on Coagulation Function and Nerve Function of Patients with Ischemic Stroke. Evidence-based complementary and alternative medicine : eCAM 2022;2022:9440271.

32. P S, R H, M T, et al. Is Unexplained Early Neurological Deterioration After Intravenous Thrombolysis Associated With Thrombus Extension? Stroke 2017;48:348–352.

33. JF B-C, M A-J, Y S. Clinical deterioration following middle cerebral artery hemodynamic changes after intravenous thrombolysis for acute ischemic stroke. Journal of stroke and cerebrovascular diseases : the official journal of National Stroke Association 2014;23:254–258.

